# Eye Movement Variations in Indoor, Outdoor, and Reading Scenarios and their Implications for Myopia

**DOI:** 10.1101/2024.07.20.24310744

**Authors:** Qi Li, Chao Zhou, Tingting Liu, Yingxiang Han, Dajiang Wang, Xiaofei Wang

**Affiliations:** Key Laboratory for Biomechanics and Mechanobiology of Ministry of Education, Beijing Advanced Innovation Center for Biomedical Engineering, School of Biological Science and Medical Engineering, Beihang University, Beijing, China; Aerospace Center Hospital, Beijing, China; Senior Department of Ophthalmology, The Third Medical Center of PLA General Hospital, Beijing, China; State Key Laboratory of Ophthalmology, Optometry and Visual Science, Eye Hospital, School of Ophthalmology and Optometry, Wenzhou Medical University, Wenzhou, China

**Keywords:** Eye Movements, Myopia, Eye Tracking, Virtual Reality, Reading

## Abstract

**Purpose:** To quantitatively measure eye movement behaviors in indoor, outdoor, and reading scenarios to understand their potential link to myopia.

**Methods:** Forty-one healthy adult subjects freely viewed indoor and outdoor scenes and performed reading activities using virtual reality (VR). Eye movement data were recorded with the built-in eye tracker of the VR headset (HTC Vive Pro Eye). Gaze and fixation data were calculated and reported for eight regions of the visual field.

**Results:** Indoor scenes exhibited a more pronounced downward gaze than outdoor environments. Significant differences (p < 0.05) in gaze and fixation behaviors were observed between reading and other scenarios. In region 8 (peripheral inferior visual field), the median (1st quartile, 3rd quartile) number of gaze points were 816 (463, 1175), 1123 (743, 1497), and 1705 (966, 2382) for outdoor, indoor and reading scenarios, respectively. Similarly, fixation behavior counts were 4 (1, 9), 7 (1, 11), and 39 (22, 54), respectively.

**Conclusions:** Downward gaze and fixation behaviors are more prevalent in reading and indoor environments. Given that downwards eye movements can induce instantaneous axial elongation, our results suggested a potential biomechanical pathway for myopia progression through optic nerve traction and ocular tissue remodeling. This study underscores the need for further research to explore the specific role of eye movement behaviors in the progression of myopia, especially in real-life settings.

## INTRODUCTION

The global prevalence of myopia, particularly in East Asia, has become a major public health issue.^1,2^ Beyond impairing vision quality, myopia can progress to pathologic myopia, leading to severe complications such as foveoschisis, retinal detachment and glaucoma.^1^ Despite extensive research, the exact mechanisms underlying myopia remain unclear.

Education and intensive near-work activities, such as reading, are major risk factors for myopia.^2^ Consistent evidences have shown that outdoor activities can effectively slow down myopia progression.^3,4^ However, the underlying mechanism are not fully understood. Current research largely focus on the role of light exposure,^5,6^ but the findings have been mixed, with some^7^ supporting the hypothesis and others presenting contradictory evidence.^8–10^ Besides light intensity, other factors might be in play in the development and progression of myopia. For example, studies have shown that reading has a greater impact on students’ myopia progression than light intensity.^11,12^

Our previous studies^13–16^ and others’^17,18^ have shown that the optic nerve traction force in the antero-posterior direction during eye movement could deform the posterior eye globe and potentially lead to growth and remodelling of ocular tissues. Consistent with these findings, adduction and downward gaze have been linked to transient axial length increases, independent of accommodation effects.^19–21^ Additionally, interventions like prism lens wear, which reduce convergence eye movement, have been shown to slow myopia progression.^22^ These findings suggest that eye movement behavior across various environments and activities may contribute to myopia progression, yet the specific patterns of these movements have not been previously studied. Therefore, the aim of this study was to quantitatively measure eye movement behaviors in indoor and outdoor environments and during reading, using eye tracking technology combined with virtual reality (VR).

## METHODS

In this study, three distinct scenarios were used to analyze eye movement characteristics: viewing indoor scenes, viewing outdoor scenes and reading. Eye movement data were recorded under these scenarios to observe variations in gaze and fixation patterns.

### Subject Recruitment and Characteristics

This study involved 41 healthy adult subjects (mean age = 24 years, standard deviation = 3 years, 24 males and 17 females). All subjects had myopia with refractive errors ranging from −1.00 to −6.00 diopters, which were measured using an autorefractor. Each subject wore corrective glasses to achieve optimal visual acuity during the study. Exclusion criteria included the presence of abnormal stereo vision, strabismus, retinal disease, visual field defects or other eye diseases, as well as restrictions in body movement and balance, difficulties in comprehending or adhering to the instructions provided by the research team, or an inability to engage in the full extent of the experimental protocol. Written informed consent was obtained from all participants. The study was approved by the Biomedical Ethics Committee of Beihang University and was conducted in accordance with the guidelines of the Declaration of Helsinki.

### Virtual Reality Setup

The VR device used in this study was HTC VIVE Pro Eye head-mounted display (HMD), featuring a built-in eye tracker provided by Tobii (Tobii AB, Stockholm, Sweden). The eye tracker offers a data output frequency of up to 120 Hz and an accuracy range of 0.5°–1.1°. Raw data of the eye tracker can be exported using the HTC SRanipal software development kit.

To simulate both indoor and outdoor scenes, DeoVR Video Player was utilized. 360° panoramic images were projected into the VR’s 3D space using Equi-Rectangular Projection. To simulate reading scenes, VR Kindle application was utilized. The e-book was displayed in 2D Panel Mode, allowing the user to adjust the location of the virtual Kindle to achieve a comfortable reading distance.

### VR Scene Selection and Categorization

VR scene images were sourced from a 360° panoramic image database compiled by Rai et al.^23^ From this collection, we selected 12 images depicting indoor spaces (6 images) and urban outdoor spaces (6 images) to represent the indoor and outdoor scenarios in our study. The resolution of the chosen images was greater than 5760×2880, facilitating the creation of clear, immersive environments. **Figure 1** displays the 360° panoramic views of the scene images for the indoor and outdoor settings.

**Figure 1.**
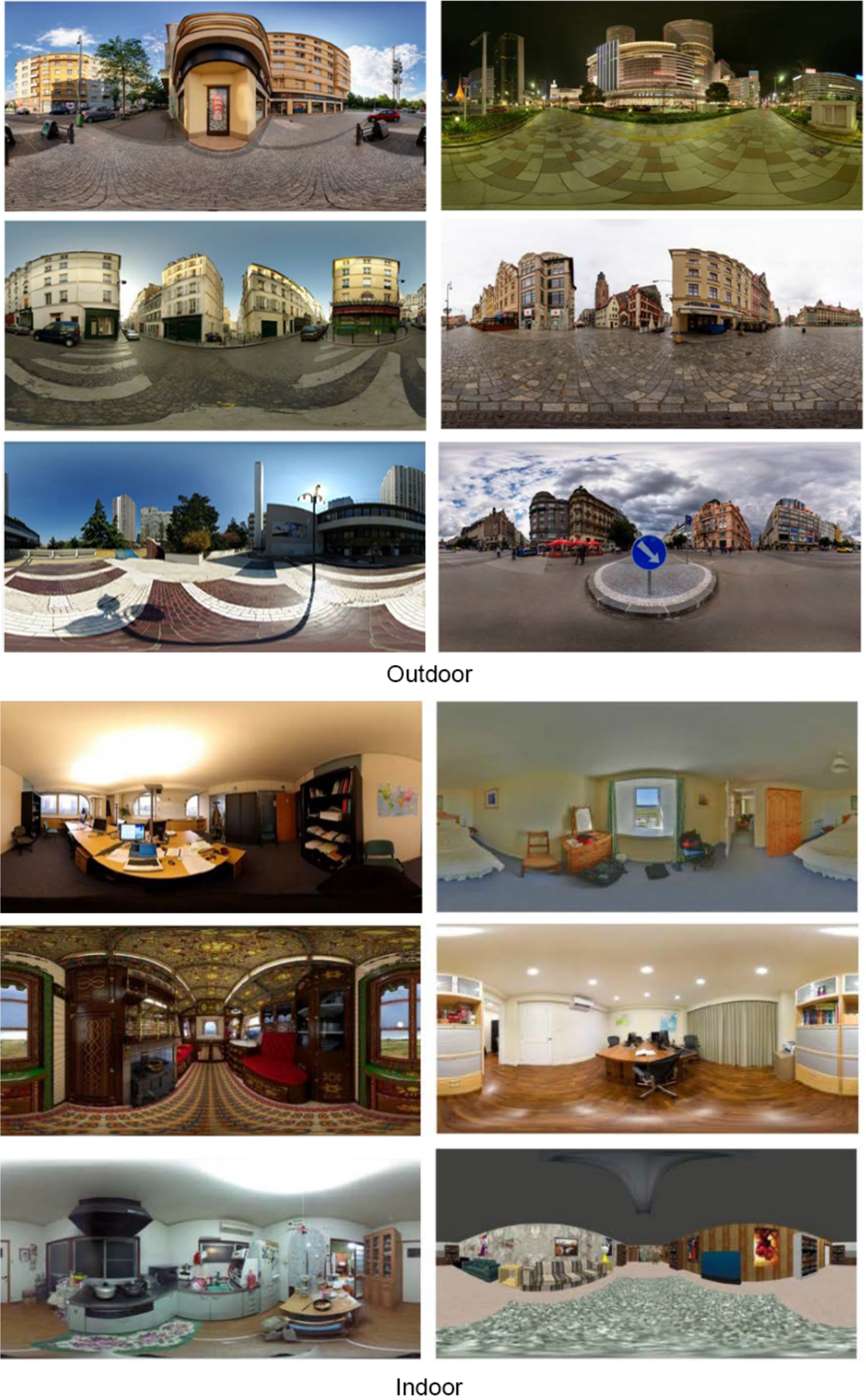
Panoramic images of the indoor and outdoor scenes used in this study. The outdoor scene primarily consists of urban open spaces and the indoor scenes feature various room spaces.

### Experimental Procedure and Data Recording

Before the experiment, each subject underwent an eye movement calibration. Following this, we launched the DeoVR scene player and data recording applications to initiate the experiment. At the start, subjects were oriented to face the center of the image by following guidance arrows to locate a target at the image’s center. They were instructed to close their eyes and rest until a countdown reached zero, at which point they opened their eyes to freely explore the VR environment. During the free exploration, no further instructions were given until the exploration ended.

Subjects viewed each image for 25 seconds. Subsequently, they were reoriented to the image’s center and closed their eyes for a 10-second rest before transitioning to the next image. After viewing all the scene images, subjects engaged in reading a book in Kindle VR for 180 seconds until the phrase “experiment ends” appeared in the VR scene.

Subjects were offered breaks after viewing each group of images to prevent symptoms of discomfort such as dizziness or nausea, which can arise from VR incompatibility. If a break was needed, they could remove the HMD and rest. Before resuming, the eye movement calibration step was repeated.

The recorded eye movement data included timestamps, gaze origin and gaze vector, data reliability, pupil size, eye openness for both eyes. Only the data from the left eye was analyzed, as the data from both eyes were similar.

### Data Processing and Eye Movement Parameters Calculation

The raw data were exported as text files and later converted to .csv files for data processing. The beginning of each viewing session was identified using the eye openness data, as subjects were instructed to close their eyes before starting tasks. Subsequently, a segment of 25 seconds of data following the beginning was extracted, representing the eye movement data for that specific image.

In the indoor and outdoor scenarios, for each scene image of a single subject, the eye movement behaviors were analyzed. The eye movement parameters were then calculated for each image of each subject. For reading, recorded data were segmented into durations of 25s and the first six segments were analyzed. This approach is similar to the analysis of the six images in the indoor and outdoor tasks.

Eye movement behavior includes two major types: fixation behavior and saccade behavior. Fixation was identified using the Dispersion Threshold Identification (I-DT) algorithm.^24^ The dispersion threshold of 1° and duration threshold of 150ms was used in the algorithm.

### Measurements of Gaze and Fixation Behaviors

Gaze behavior was represented through gaze points in the visual field. Specifically, gaze points were obtained by projecting the gaze vector onto a spherical surface centered on the eye.^25^ These gaze points on the spherical surface were then plotted as a 2D map, similar to a standard human visual field map. The visual field was divided into four sectors: nasal, superior, temporal, and inferior. Each sector was further divided into two subregions, with a 10° central vision boundary line as shown in **Figure 2**.

**Figure 2.**
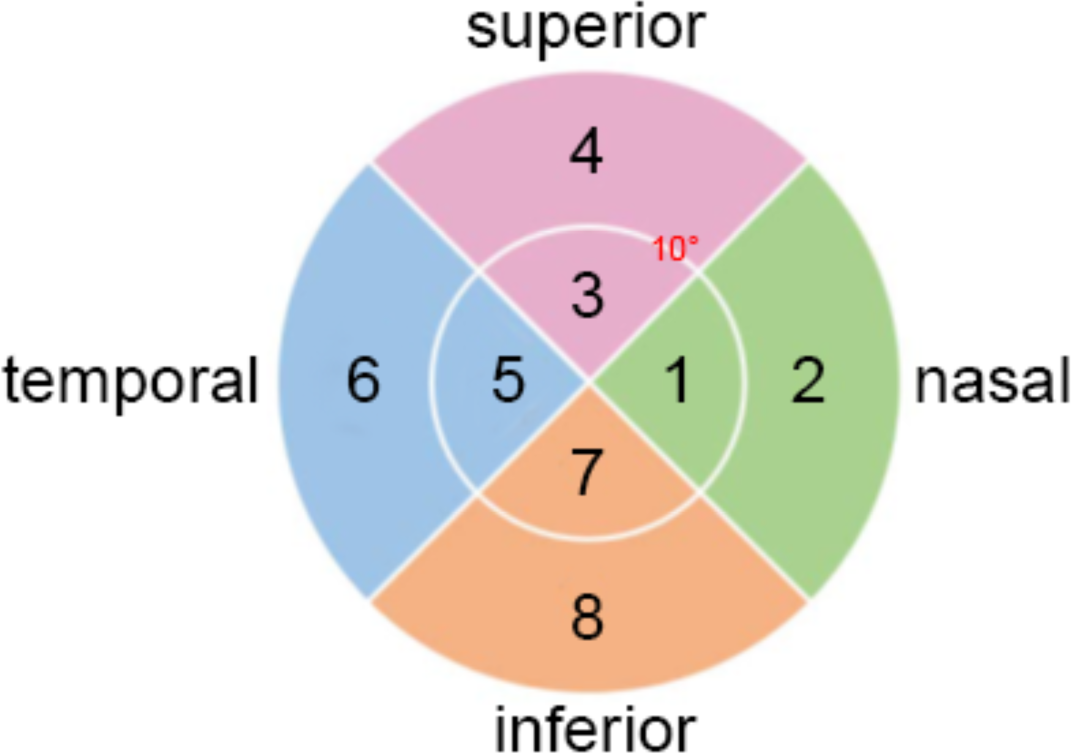
Illustration of the 8 regions of the visual field used in this study. The visual field was divided into four sectors: nasal, superior, temporal, and inferior. Each sector was further divided into two subregions, with a 10° boundary for central vision.

The density of gaze points on the visual field were plotted using a probability density map.^26,27^ High density area indicates where subjects gaze more frequently, as eye position data were recorded at a constant frequency. For each scene within the indoor and outdoor scenarios, gaze points for each subject were analyzed.

The gaze points or region-wise gaze points for each individual subject were calculated separately for all 6 scenes in both indoor and outdoor scenarios. For reading, gaze points were calculated for each subject during six consecutive 25s periods to align with the six images per scenario in both indoor and outdoor scenarios. ensuring consistent duration and data segments across all conditions.

For fixation behavior, individual fixation locations in the visual field and fixation durations were obtained using the I-DT algorithm. These fixation points were then plotted as a probability density map, similar to the gaze behavior analysis. Subsequently, the number of gaze events at each sector for a specific subject across different scenarios was calculated, mirroring the approach used for gaze behavior analysis.

### Statistical Analysis

The Kolmogorov-Smirnov test was used to compare the differences in gaze and fixation behaviors between different scenarios. As multiple comparisons were made, Bonferroni corrections were applied to adjust the p-values for significance. A p-value of less than 0.05 was considered statistically significant.

## RESULTS

### Gaze Behavior

The probability density map in **Figure 3** illustrates the gaze behavior across three scenarios: viewing indoor scenes, viewing outdoor scenes and reading. The probability density of gaze points in reading (maximum probability density of around 19) is much higher than that in indoor (maximum probability density of around 6) and outdoor (maximum probability density of around 7). Gaze points were largely concentrated in the inferior regions (region 7 and 8) for all three scenarios.

**Figure 3.**
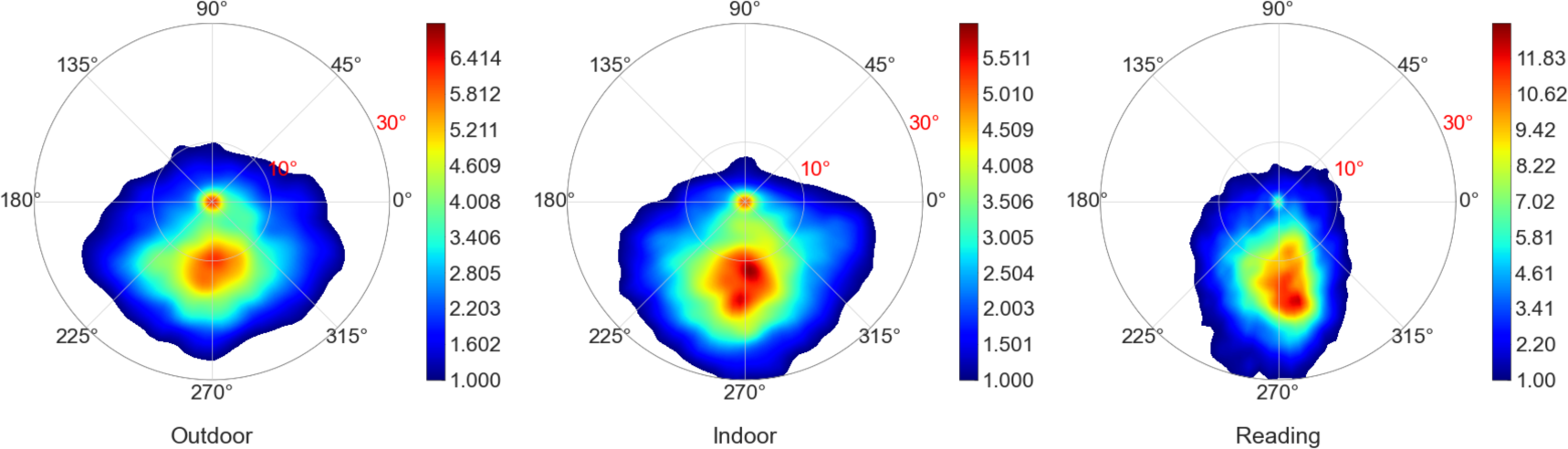
Gaze points probability density map on the visual field in all three scenarios. Note that the color scale ranges in each subfigure vary to better illustrate the relative distribution within each scenario.

Further, specific the number of gaze points in different regions for all subjects in each scenario were shown in **Figure 4**. There were significant differences between the reading and the other two scenarios in all 8 regions. The number of gaze points in region 8 is also listed in **Table 1**. The number of gaze points in region 8 was highest during reading, followed by the indoor scenario, and then the outdoor scenario, suggesting more frequent eye movement towards the inferior direction in reading and indoor scenarios.

**Figure 4.**
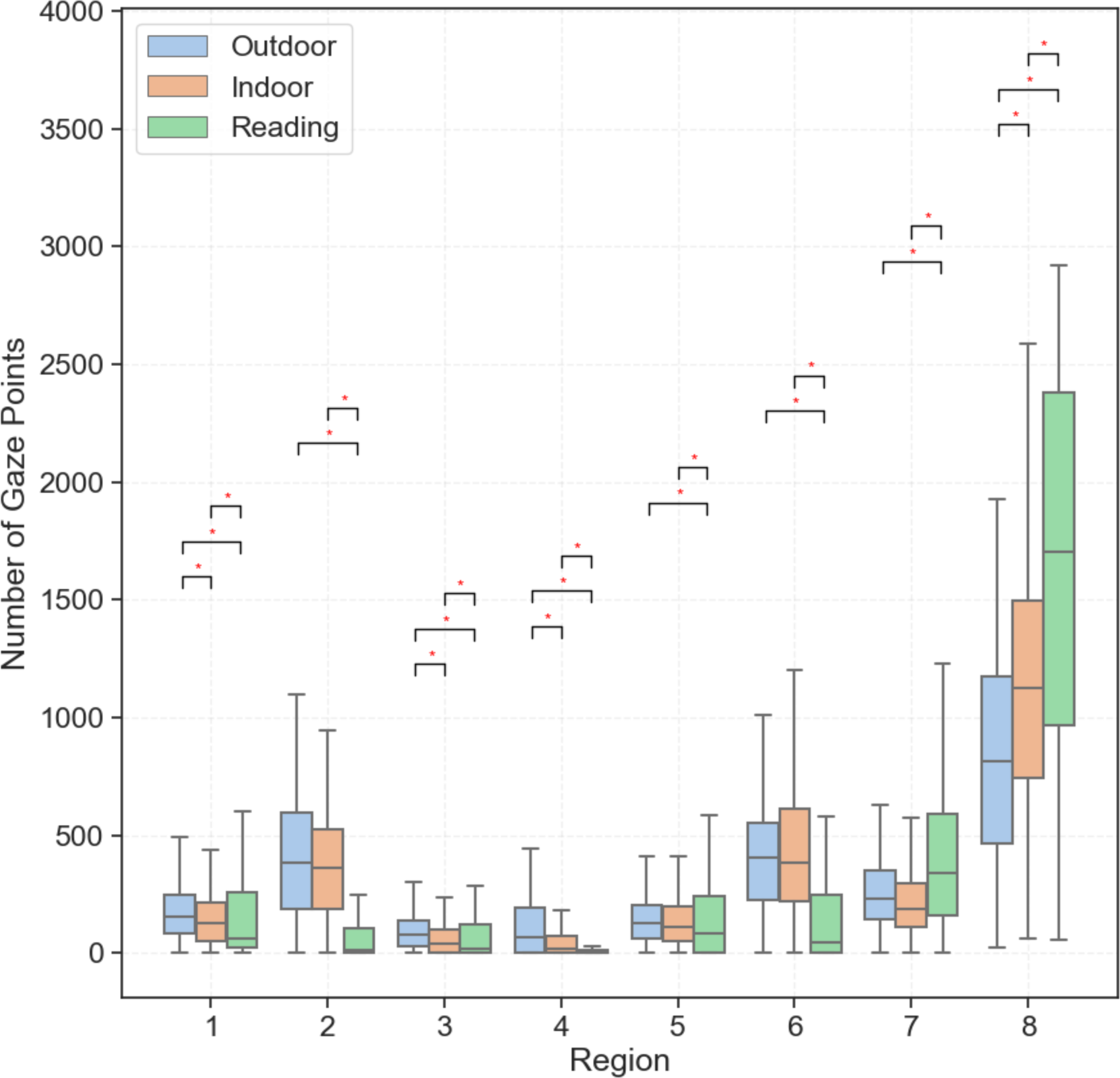
Box plot of the number of gaze points in different regions across all three scenarios. * indicates statistically significant differences.

**Table 1.** Number of gaze points and fixation times in region 8.

| Scenarios | Number of gaze points | Number of fixation times |
| --- | --- | --- |
|  | Median (first quartile, third quartile) | Median (first quartile, third quartile) |
| Outdoor | 816 (463, 1175) | 4 (1, 9) |
| Indoor | 1123 (743, 1497) | 7 (2, 11) |
| Reading | 1705 (966, 2382) | 39 (22, 54) |

The number of gaze points on nasal and temporal sides (region 2 and 6) in reading is obviously less than that in indoor and outdoor. That indicates that the majority of the gaze points were within a relatively narrow region in reading compared to indoor and outdoor scenarios.

### Fixation Behavior

**Figure 5** shows the probability density of fixation points. **Figure 6** shows the number of fixation times in different regions for all subjects across the three scenarios. There were significant differences between the reading scenario and the other two scenarios in region 1, 5, 7 and 8. The number of fixations in region 8 is shown in **Table 1**. There was also a significantly higher number of fixations in the reading and indoor scenario than in the outdoor scenario.

**Figure 5.**
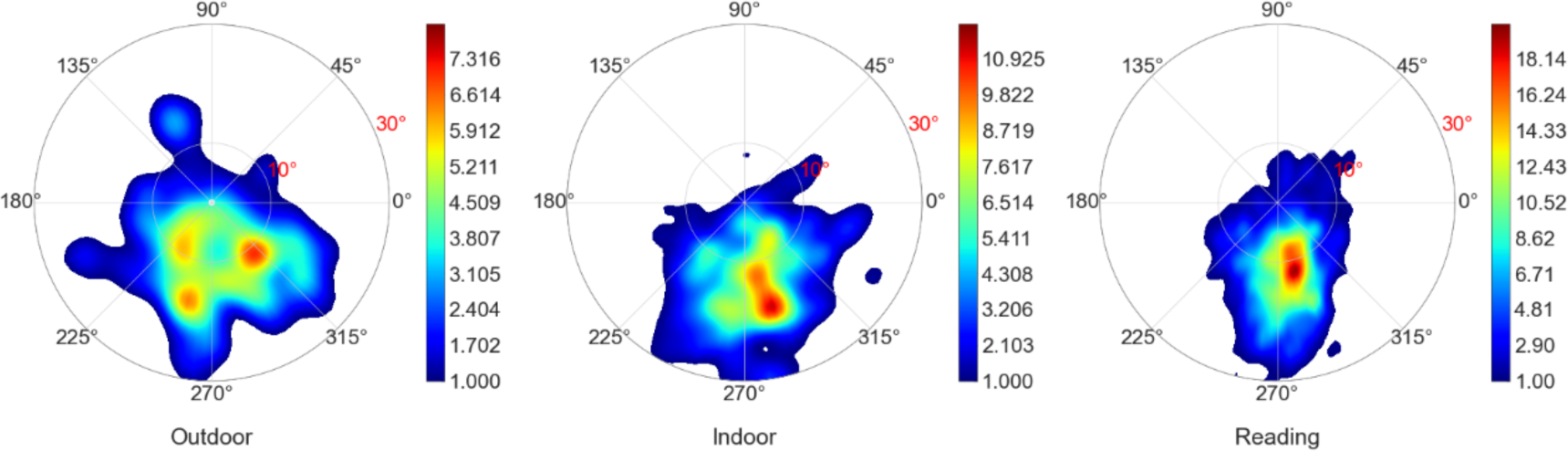
Fixation points probability density map on the visual field in all three scenarios. Note that the color scale ranges in each subfigure vary to better illustrate the relative distribution within each scenario.

**Figure 6.**
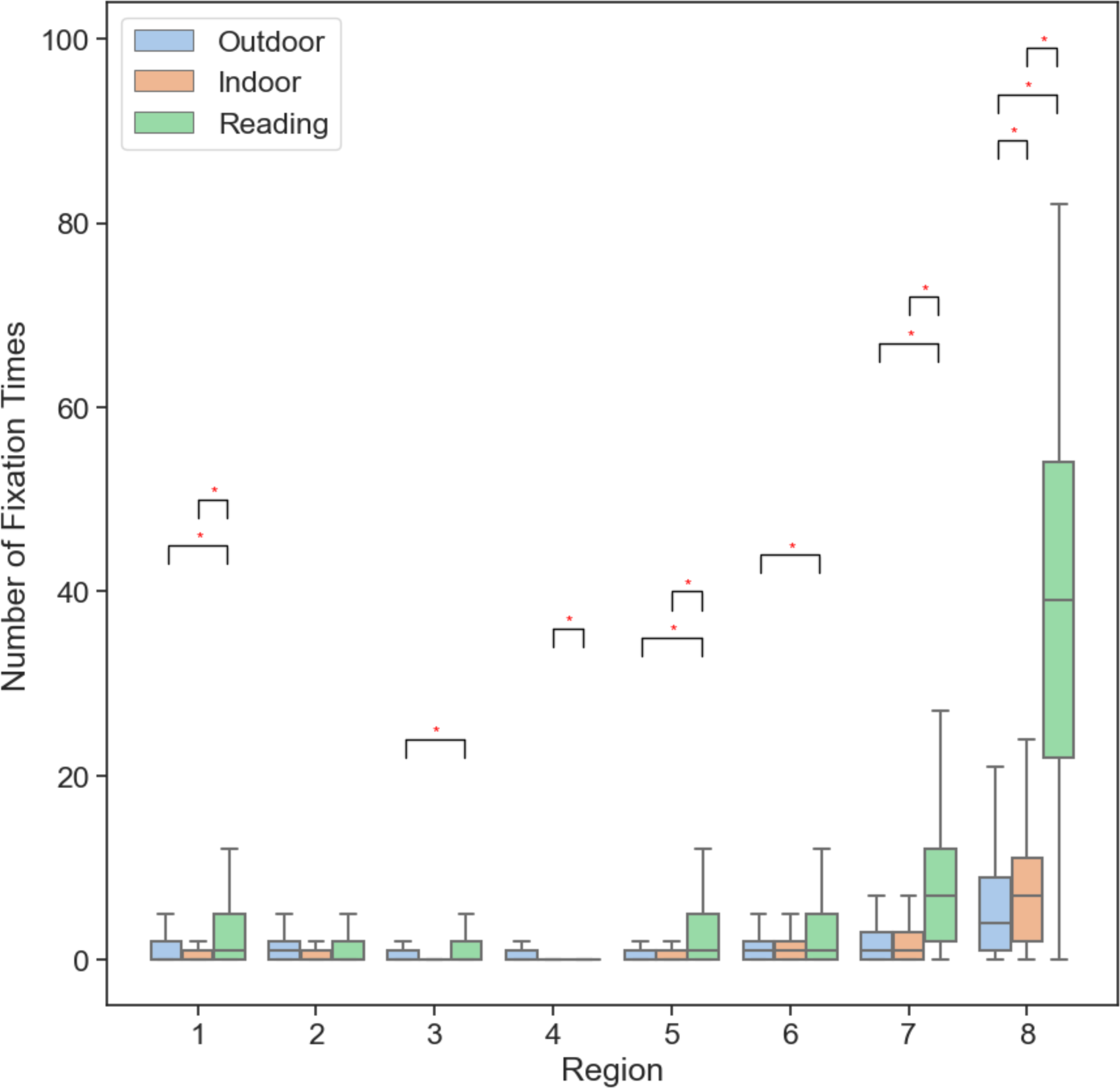
Box plot of the number of fixation times in different regions across all three scenarios. * Indicates statistically significant differences.

## DISCUSSION

In this study, we found distinct eye movement behaviors across various scenarios, with a notably higher frequency of downward gaze and fixation during reading compared to both indoor and outdoor settings. Indoor environments also exhibited more downward gaze and fixation than outdoor ones. According to previous studies^13–18^, downward eye movement has the potential to cause transient axial elongation and could potentially have longitudinal effects.

Our findings highlight a fundamental difference in the eye movement patterns associated with reading, compared to those observed in outdoor and indoor environments. Inferior areas consistently showed more gaze points and fixation behaviors across all scenarios, particularly in reading. Notably, both indoor and reading scenarios showed a higher count of these points compared to outdoor scenarios.

Reading is a well-known risk factor for myopia. Our data showed that reading involves more downwards gaze and potentially larger accumulative optic nerve pulling in the antero-posterior direction, while outdoor activity has the least downward eye movement. Our previous study^14^ has shown that the traction force of the optic nerve on the eyeball during eye movement is comparable to the force exerted by extraocular muscles and acts in the antero-posterior direction. This traction force can deform the optic nerve head and the deformation can be larger than those induced by a substantial intraocular pressure elevation.^13,15^ Optic nerve head deformations induced by eye movement has been shown to be correlated with normal tension glaucoma.^28^ Growth and remodeling in tissues are influenced by mechanical factors such as stress, leading to the concept of “stress-modulated growth”.^29^ The optic nerve traction force, if sustained or repeated over a long period, could potentially stimulate the growth and remodeling of the eyeball. This assumption aligned well with previous studies which have shown that downward eye movements (without accommodation) are associated with a temporary increase in axial length.^19–21,30^ Additionally, wearing prism lens to reduce eye convergence during near work can slow myopia development.^22^ Furthermore, in the guinea pig myopia model, eye elongation initially occurred around the optic nerve rather than the macula.^31^ These findings suggest that the optic nerve traction could be a factor related to myopia. This study provides some indirect evidences and future studies with longitudinal data are needed to confirm this hypothesis.

Time spent outdoors is known to be protective against myopia, but the underlying mechanism remains elusive. Previous studies have primarily focused on the role of light exposure during outdoor time,^5,6^ with some suggesting that moderately bright light increases retinal dopamine levels, thereby offering some protection against myopia.^7^ However, there are also studies that challenged this hypothesis,^8,9^ which argued that light may not be the actual mechanism.^10^ The spatial frequency characteristics of difference scenes may be one of the missing drivers of the form deprivation myopia.^32^ Our results suggested that eye movement behaviors differ during outdoor activity compared to indoor and near work, such as reading. It’s a new hypothesis that these differences in eye movement behavior may be one factor contributing to the effectiveness of outdoor activities in mitigating myopia progression. Further research is warranted to investigate the role of eye movement in outdoor activities and its potential correlation with myopia progression.

Previous studies on eye movement behaviors in different scenes have primarily focused on the restorative effects, i.e., the recovery from mental fatigue and stress, and their impact on the physiological and psychological conditions of observers in natural scenes compared to urban and modern architecture.^33–36^ These studies focused on visual attention towards targets, not the eyeball. In contrast, this study examines the eyeball’s movement behaviors across different scenarios. Furthermore, we used a head-mounted eye tracker to eliminate the effects of head movement, allowing us to measure eye movement behaviors while subjects freely moved their heads and eyes. This approach is different from other methods that restrict head movement to a small range. Moreover, we employed VR technology to conduct the experiments. Previous studies have shown that 3D cues are fundamentally different from those in 2D pictures, which only provide intensity, texture, and color cues.^37^ The VR technology allowed subjects to observe the scene in a more immersive manner, differing from other eye movement studies that used a laptop screen to display a scene and asked the subject to view it from an aerial perspective, which is far from reality.

In this study, especially in the indoor scenario, subjects were asked to explore freely in the room. However, this is not typical behavior for most people during indoor activities, which usually involve specific tasks. For example, students usually remain indoors for academic activities such as attending classes and studying. At home, they might engage in completing homework, reading, or preparing for exams, along with other indoor leisure and screen time. Therefore, real-life indoor scenarios often involve more near work compared to the tasks in this study, likely resulting in a more pronounced downward gaze due to these activities. However, even for free viewing tasks, subjects still move their eyes more in the downward direction due to the region of interest or object of interest being more concentrated below eye level, showing more downward gaze and fixation behaviors. Further studies that accurately replicate everyday indoor activities will enhance our understanding of eye movement behaviors in indoor spaces.

We used gaze and fixation patterns to quantify eye movement behaviors in this study. Prior studies have indicated that greater eye movement magnitude is associated with increased traction force of the optic nerve,^38^ thereby exerting more force on the posterior sclera of the eyeball. However, the precise influence of force duration at a specific magnitude remains unclear. It is plausible that brief, instantaneous forces, such as those experienced during saccades, might have a lesser effect on tissue growth and remodeling compared to a prolonged period of sustained force of the same magnitude and cumulative duration during fixation. Elucidating this requires a fundamental understanding of scleral growth and remodeling processes. Further investigations into the sclera’s response to varying force magnitudes and durations are warranted to clarify this point.

## Conclusion

This study identified unique patterns of eye movement across different scenarios, with a notably higher frequency of downward gaze and fixation during reading and indoor scenarios compared to outdoor settings. Such persistent downward eye movements may be associated with an increased risk of developing myopia, suggesting a potential link that warrants further investigation.

## Data Availability

All data produced in the present study are available upon reasonable request to the authors

## ACKNOWLEDGMENTS

This research was supported by National Natural Science Foundation of China (12272030, 12002025), Capital’s Funds for Health Improvement and Research (2022-2-5041), Beijing Natural Science Foundation (Z240017) and the Fundamental Research Funds for the Central Universities.

